# Adjusting for medication status in genome-wide association studies

**DOI:** 10.1101/2024.02.19.24303028

**Authors:** Amanda H.W. Chong, Christopher Kintu, Yoonsu Cho, Segun Fatumo, Jason Torres, George Davey Smith, Tom R. Gaunt, Gibran Hemani

## Abstract

When conducting genome-wide association studies, addressing medication status that is relevant to the trait of interest can prevent biases that could distort estimates of the true genetic effect. Here, we introduce the genetic empirical medication reduction adjustment (GEMRA) method which uses a heuristic search for an empirical adjustment to be applied to phenotypic values of participants reporting medication use. Through simulations we show that the direct genetic effect estimates in the GEMRA approach exhibited less bias and greater statistical power than common strategies such as including medication as a covariate, restricting the sample to unmedicated users or including all samples without adjustment. We then applied the GEMRA approach to estimate statin medication adjustment for analysis of low-density lipoprotein (LDL) cholesterol levels and antihypertensive medication adjustment for analyses of systolic (SBP) and diastolic (DBP) blood pressure using multi-population data from UK Biobank and the Uganda Genome Resource. We found that a relative rather than an absolute adjustment better modelled the effect of medication on LDL cholesterol, SBP and DBP, with an effect of 40%, 10% and 10% reduction appearing to be consistent across populations, respectively. We also show that spurious differences in medication effects across populations can arise when the effect scale is misspecified.

**Author Summary:** When studying how genetics influence health outcomes, it is important to account for medications that can affect the trait being studied. Otherwise, medication use may make genetic effects appear stronger or weaker than they truly are. Here, we developed a method, GEMRA, to account for the effects of medication in genetic studies. Using simulated data, we found that GEMRA was better able to detect true genetic associations than several commonly used approaches. We then used GEMRA to account for the effects of statins on LDL cholesterol and blood pressure medications on systolic and diastolic blood pressure. Using data from UK Biobank and the Ugandan Genome Resource, we found that medication effects were better represented as a percentage reduction rather than as the same fixed change for everyone. Our findings also showed that choosing the wrong way to represent medication effects (i.e. as a percentage reduction or fixed value) can create misleading differences between populations.

## Introduction

Inclusion of participants taking medication in genome-wide association studies (GWAS) of a phenotype affected by that medication is challenging as it introduces heterogeneity in the analyses. This is due to medicated participants having an observed phenotype value that is not reflective of their true, unmedicated phenotype value included in the analyses with unmedicated participants, resulting in findings that don’t represent either group (1–3). For example, participants taking LDL-lowering medication will have a lower observed LDL cholesterol (LDLc) value due to the effect of medication use. This issue has been previously explored by Bell et al. 2022(4) and Fang et al. 2022(5), with both findings showing the misleading apparent effects of higher BMI on lowering LDLc when medication status is not addressed. However, when medication use is considered, these results can be attributable to people with higher body mass index (BMI) being more likely to be given statins, a LDL-lowering medication.

To date, there is no approach to correct for medication status that would result in an adjustment that reliably restores the medication-adjusted phenotype value to the unmedicated value. In previous literature, approximations of the unmedicated phenotypic value have been applied for phenotypes such as blood pressure(6–8), lipid traits(9–12), and type 2 diabetes(7, 9, 13) with studies accounting for treatment effect using a plausible constant, censored normal regression, or a non-parametric method(6, 14). However, studies have commonly excluded participants based on medication status(15–22), and there are several concerns that can occur due to this approach. As shown in Figure 1, in the example involving LDL-lowering medication and LDLc, the estimand of principle interest in GWAS is typically the direct effect of a genetic variant influencing the target phenotype, sometimes described as the effect in the participant of inheriting an allele(23). However the exclusion or inclusion of participants conditioning based on medication status or stratifying by medication status opens a causal pathway through medication status to the observed phenotype value (i.e., indirect effect (*b****_y_***)). This can introduce bias to the estimated causal effect (i.e. collider bias)(24). Furthermore, additional indirect effects induced by collider bias can also be introduced as prescription of medication may be influenced by other factors other than high LDLc such as BMI, cardiovascular event, or a family history of high cholesterol (i.e., genetic variants associated with other risk factors in Figure 1). Therefore, selection by medication status could induce an association between genetic variants associated with non-LDLc risk factors and LDLc. In addition, by excluding participants by medication status, this approach could be ignoring participants that could better inform analyses of potential genetic variants associated with dyslipidemia(25). Exclusion would also decrease the sample size, reducing the statistical power of the GWAS which, in especially samples of older individuals such as UK Biobank, could be substantial(26). In contrast, not adjusting for medication status would introduce heterogeneity into the analyses as the observed value would be different from the unmedicated value. This could then result in a bias that either overestimates or underestimates the true effect, or opens a path for genetic variants affecting other factors influencing medication use.

**Figure 1.**
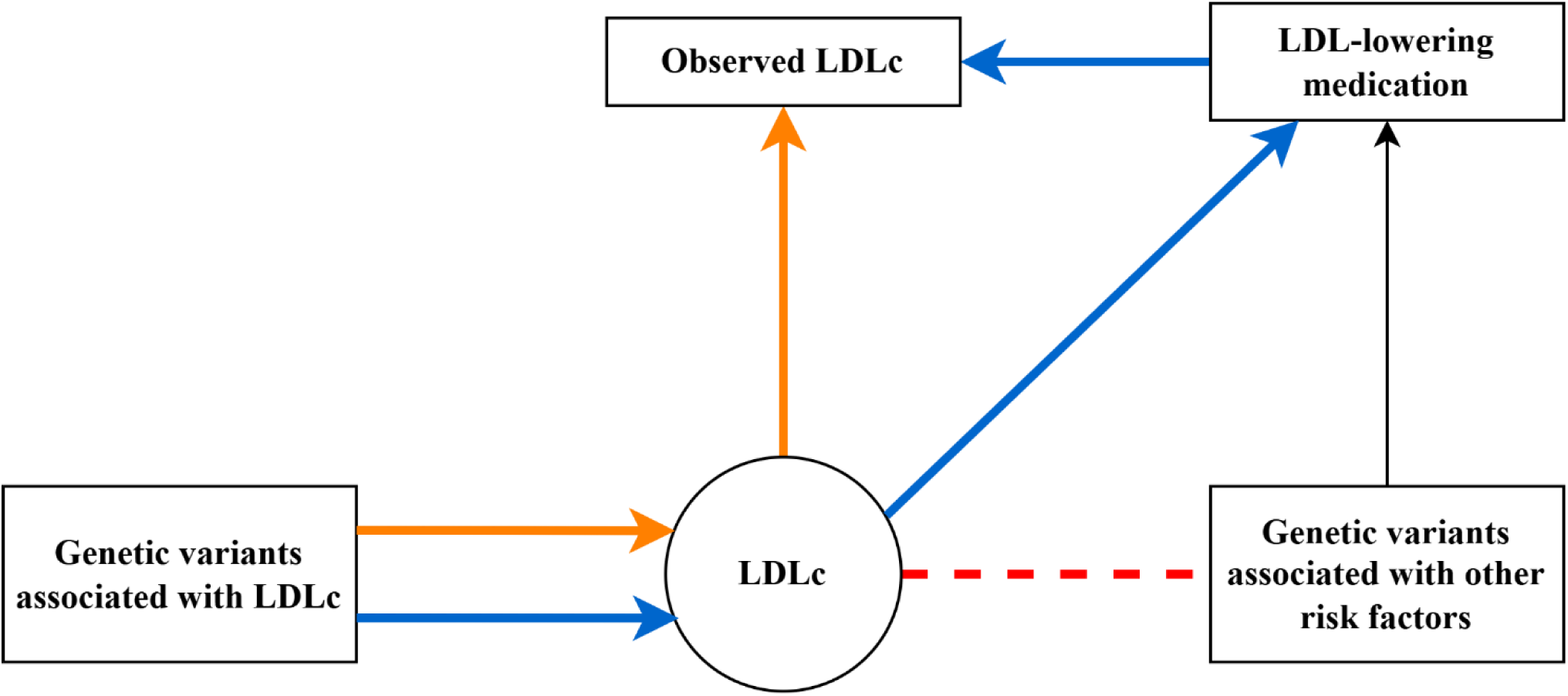
Directed acyclic graph (DAG) illustrating the direct effect of genetic variants associated with LDLc have on the observed LDLc (orange), the indirect effect (blue) of how selection of samples based on LDL-lowering medication use can introduce collider bias (indicated by the dashed red line), as conditioning on or adjusting for medication use can induce an association between non-LDLc genetic factors and LDLc. This is because non-genetic factors that influence LDLc will become related to genetic variants when adjusted and stratified for these non-genetic factors or medication status. Similarly, using a complete sample, including both users and non-users of medication, effectively introduces the same collider bias path by lowering LDLc levels amongst medication users only.

Therefore, to account for medication status, a relative or absolute adjustment of medication reduction could be applied with the aim of restoring the observed phenotypic value to approximate the counterfactual value had the participant not taken medication. We propose the genetic empirical medication reduction adjustment (GEMRA) method that would heuristically search for an optimal adjustment value to approximately correct the medication-biased phenotype measurements to a value that better reflects the unmedicated phenotypic value in participants taking medication. *Relative adjustment* would be a percentage reduction in medication effect that is applied to the observed value. *Absolute adjustment* would be an additive medication reduction value that is applied to the observed value. In a cross-population setting, differences in prevalence of medication use and phenotype distribution may contribute to differences in the optimum adjustment for each population.

In this study, we applied the GEMRA approach to LDLc and LDL-lowering medication status, as well as blood pressure traits and antihypertensive medication status. We first employed a simulation framework to examine the impact of this approach on direct and indirect genetic associations, and then examined the robustness of this approach under different conditions. We then conducted the analyses using real-world data from UK Biobank (UKB) and the Uganda Genome Resource (UGR) to evaluate the adjustment approach in different populations for LDLc and blood pressure measures.

## Results

### Demonstration of collider bias issues due to medication use

Following the model outlined in Figure 1, we conducted simulations to evaluate the extent of bias on both direct and indirect effects that can arise under different GWAS strategies that attempt to manage medication status. We evaluated the bias at direct and indirect genetic effects for four strategies for handling medication status in GWAS compared to the true genetic effect: 1) Medication status is unaccounted for (1) Uncontrolled; 2) Restricted to individuals taking medication (Medication-only); 3) Restricted to individuals not taking medication (Medication-excluded); 4) Medication status included as a covariate (Medication-covariate); 5) Medication use is adjusted for (Approximate adjustment for medication)).

For the direct effects, we observed stronger attenuated genetic effect estimates for the medication-only GWAS analysis, medication-covariate and uncontrolled methods compared to the methods adjusting for medication and excluding medication status (Figure 2). The genetic effect estimates for the methods restricted to individuals not taking medication (i.e. Medication-excluded) and adjusting for medication status were strengthened and closer to the true effect estimates (Figure 2). Statistical power, assessed as the percentage difference in mean F-statistic for each method compared to the mean F-statistic for the ‘True effect’ model, was then calculated and findings showed the greatest power for the approximate adjustment for medication approach (0.9%). This was then followed by the medication-covariate (1.8%), uncontrolled, (5.8%), medication excluded (17.6%) and medication-only approaches (83.2%).

**Figure 2.**
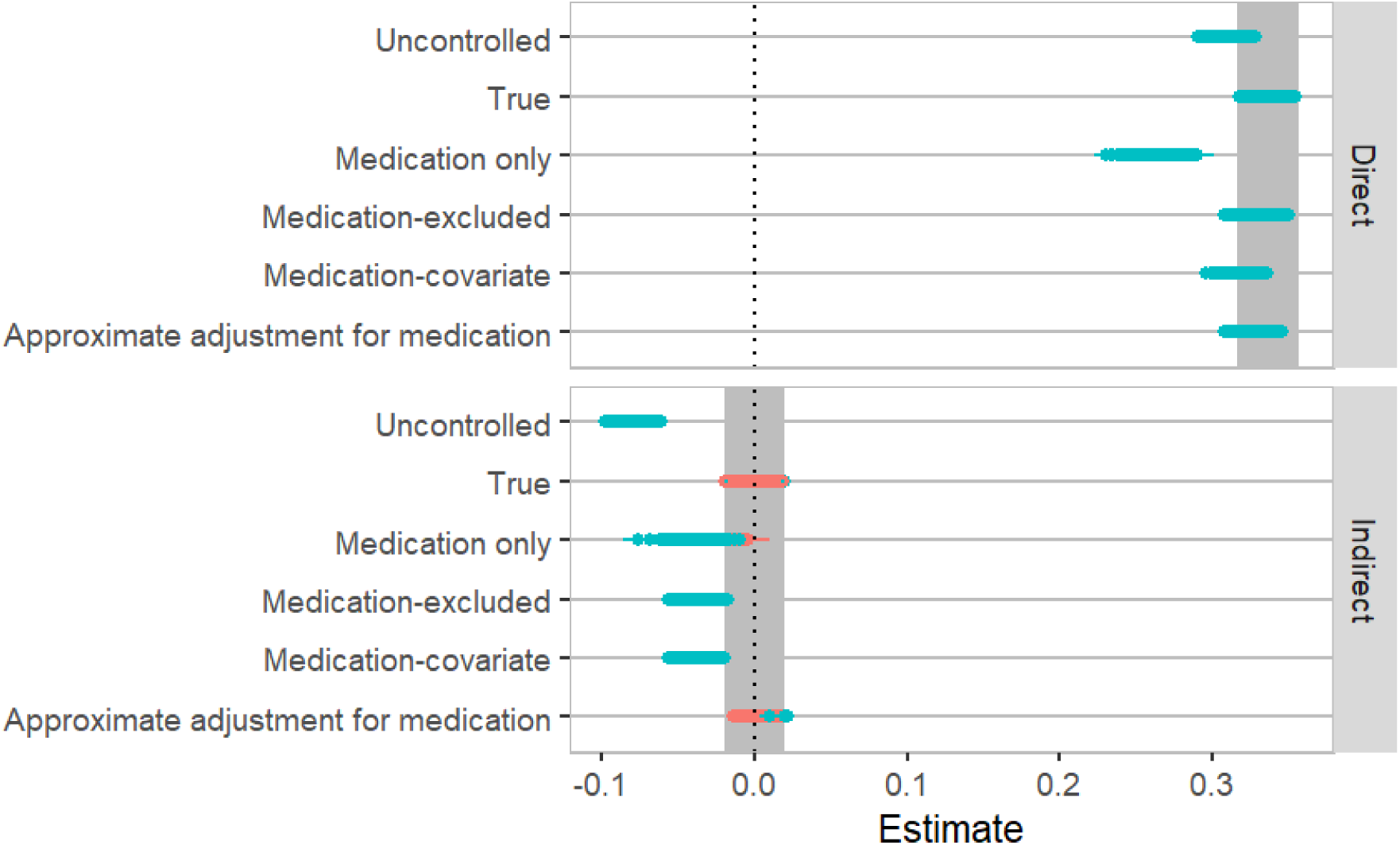
A comparison of the true genetic effect estimate precisely adjusting for medication status compared to the genetic effect estimates of the direct and indirect genotype-LDLc association when: (1) Medication status is not accounted for (Uncontrolled); (2) Restricted to individuals taking medication (Medication-only); (3) Restricted to individuals without medication (Medication excluded); (4) Medication status as a covariate; (5) Medication use is adjusted for (Approximate adjustment for medication). The blue points denote a *p*-value threshold < 0.01 indicating false-positive results and the red points indicate a *p*-value threshold > 0.01 indicating no evidence of false-positive results.

For the indirect effects, the method to approximately adjust for medication showed low false positive rate (*p* > 0.01) with a false positive rate of 0.08 similar to the true effect estimate results in comparison to the other methods with false positive rates of one for uncontrolled, 0.97 for medication only, one for medication fitted as a covariate, and one for medication excluded (Figure 2). In addition, we observed that collider bias induced a negative association between the unadjusted LDLc phenotype and non-LDLc genetic factors for the uncontrolled, medication-only, medication-covariate and medication-excluded methods.

We next evaluated how sensitive the bias was in the adjustment method to inaccuracy of the correct level of adjustment. We showed that if the medication effect is adjusted incorrectly this can still lead to bias of the genetic effect estimates on medication association *(b̂_y_*) and the genetic effect estimates on the observed exposure variable association (*β̂_y_*), though the majority of adjustment values reduced bias in comparison to the approach that performed no adjustment (Supplementary figure 1). This illustrates the importance of being able to estimate the appropriate adjustment factor in order to minimise bias. The bias impacting *b̂_y_* and *β̂_y_* was not strongly affected under the condition that medication effects were heterogeneous across individuals, although as effect heterogeneity increased so too did standard errors of the genetic effect estimate (Supplementary figure 2).

The GEMRA approach operates by seeking the medication adjustment value that maximises the variance in the phenotype explained by genetic factors for the trait of interest (*R*^2^). Supplementary figure 3 illustrates how, for a medication effect acting on the relative scale, searching across a range of relative and absolute adjustments will lead to the *R*^2^ being maximised for the true relative medication effect. However, applying either absolute or relative medication adjustments showed improvements in *R*^2^ over the unadjusted model. Secondly, when we conducted 1,000 repeats of these simulations, our results showed that the *R*^2^ in the GEMRA approach was maximised for a relative adjustment model 84% of the time compared to an absolute adjustment model, indicating that it is effective at selecting the correct effect scale (Supplementary figure 4). When we compared the mean squared error of the effect estimate using the unadjusted exposure variable against the adjusted exposure variable (i.e. adjusted on the medication parameter that maximises *R*^2^), we showed that medication adjustment always outperformed the unadjusted model even when in 16% of cases the medication adjustment parameter applied the absolute rather than the relative model (Supplementary figure 5). We also showed overall that the medication adjustment approach of the GEMRA method outperformed the unadjusted analysis even when we used the unadjusted exposure variable with medication fitted as a covariate (Supplementary figure 5). Lastly, we evaluated the issue of data leakage by comparing the direct genetic effect estimates using the GEMRA approach (i.e. Approximate adjustment for medication) and the ‘True effect’ model, and we found no evidence of bias due to data leakage with findings showing a strong, positive linear correlation (Supplementary figure 6).

### Empirical analysis: UK Biobank and Ugandan Genome Resource

We then applied these approaches to real-world data from two data sources (UKB and UGR) to evaluate the best model for LDLc and statin use, as well as blood pressure traits (i.e. SBP and DBP) and antihypertensive drug use, in each population. This was determined by the highest summed R^2^ value of GWAS hits when applying different absolute and relative parameters to adjust for medication reduced in the observed phenotype values of interest. Descriptive characteristics of LDLc, blood pressure traits and medication use in UKB across population and UGR are shown in Table 1.

**Table 1.** Description of LDL cholesterol, blood pressure traits and medication use across different populations in UK Biobank and Ugandan Genome Resource.

| Data source | Population | LDLc<br>(Median (IQR)) mmol/L | Statin use<br>(Case/controls; %) | SBP<br>(Median (IQR))<br>mmHg | DBP<br>(Median (IQR))<br>mmHg | Antihypertensive use<br>(Case/controls; %) |
| --- | --- | --- | --- | --- | --- | --- |
| UKB | African | 3.2 (2.7-3.8) | 573/3,231; 18% | 141 (126-157) | 86 (78-96) | 854/2,948; 29% |
|  | East Asian | 3.4 (2.9-3.9) | 171/1,415; 12% | 131 (117-150) | 81 (73-90) | 156/1,429; 11% |
|  | European | 3.5 (2.9-4.1) | 31,547/219,151; 14% | 136 (123-152) | 81 (74-89) | 25,155/225,471; 11% |
|  | South Asian | 3.3 (2.7-3.9) | 273/1,057; 26% | 136 (123-152) | 83 (76-91) | 156/1,174; 13% |
| UGR | Ugandan | 2.0 (1.5-2.5) | 24/6,144; 0.4% | 120 (112-130) | 73 (68-80) | 261/5,907; 4% |

For the relative medication adjustment approach, LDLc findings were consistent amongst all populations with the highest summed R^2^ value at the threshold adjusting for either 30% or 40% reduction in observed LDL due to medication (Figure 3). For SBP, findings were consistent with all populations showing the relative adjustment of 10% reduction in medication to be the best model (Figure 4). Similarly, findings for DBP showed across populations that the 10% threshold had the highest summed R^2^ value (Figure 4). Overall, the distribution in summed R^2^ values for all phenotypes were similar across populations.

**Figure 3.**
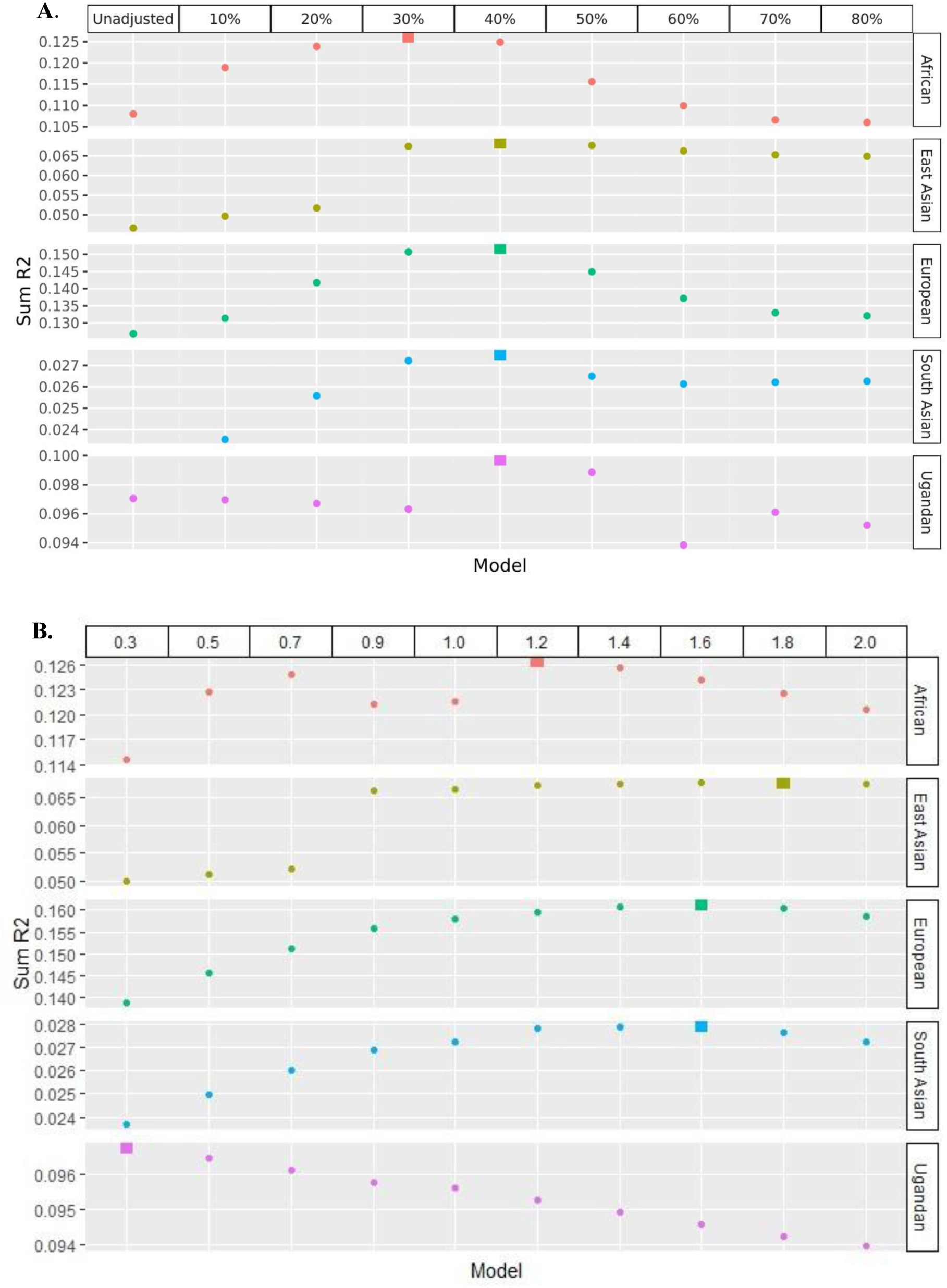
Comparison of relative (A) and absolute (B) adjustment of statins (mmol/L) in different populations using the GEMRA approach. In UKB (i.e., African, East Asian, European, and South Asian) and UGR (i.e., Ugandan), for each population, the points represent the summed R^2^ value for each model, with the large square highlighting the model with the highest summed R^2^ value.

**Figure 4.**
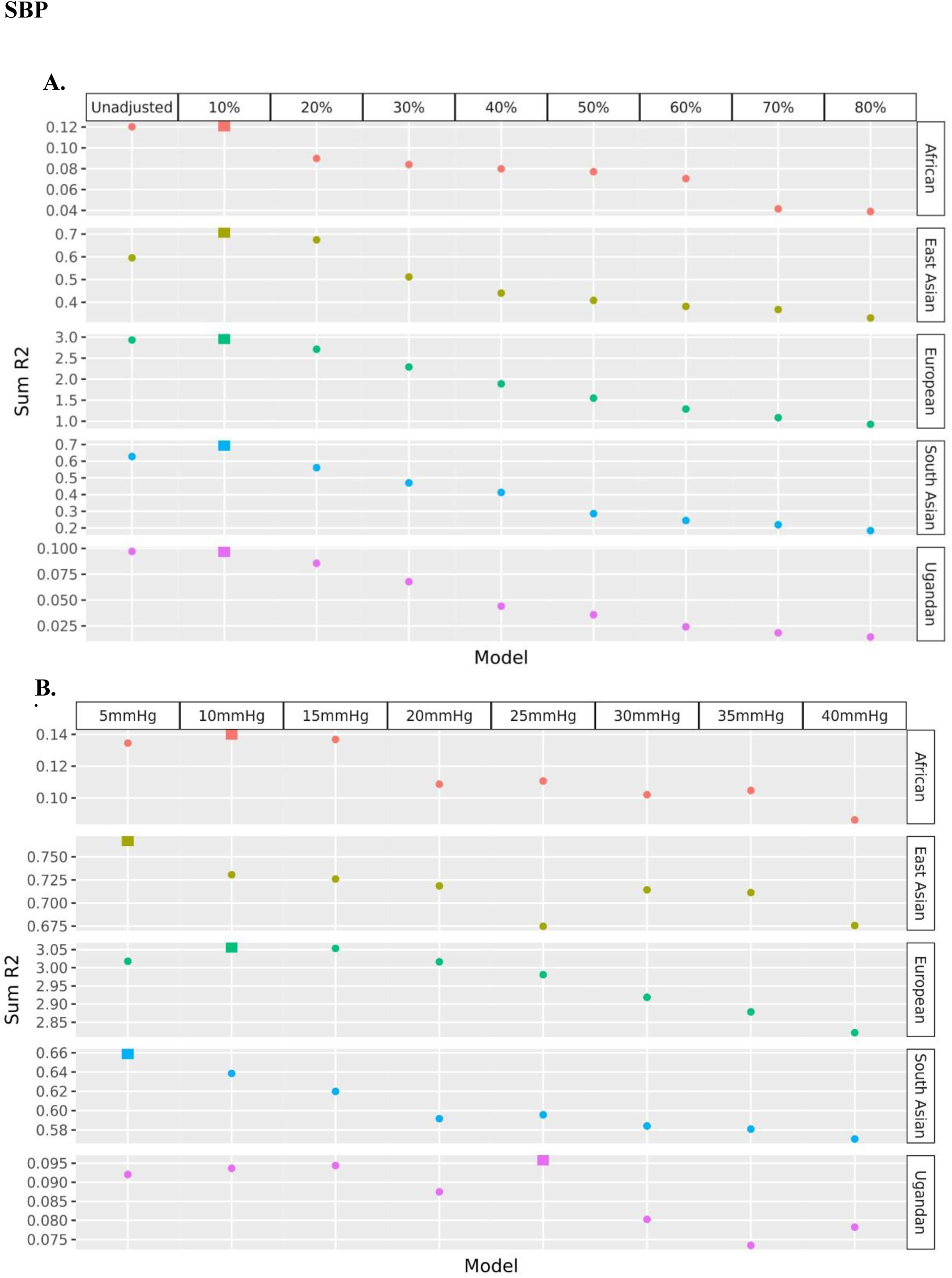
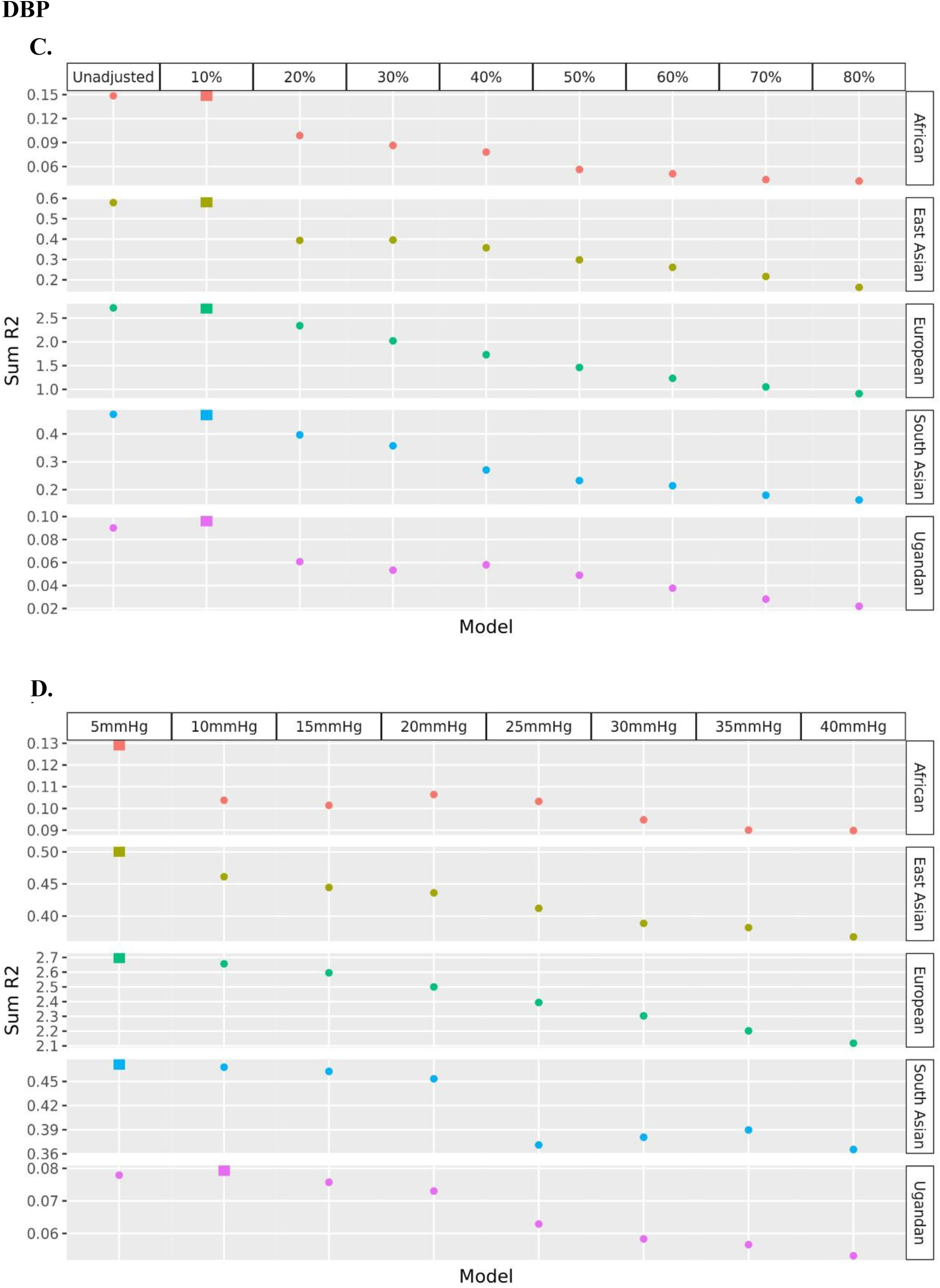
Comparison of relative and absolute adjustment of antihypertensive medication (mmHg) on SBP (A, relative; B, absolute) and DBP (C, relative; D, absolute) in different populations using the GEMRA approach. In UKB (i.e., African, East Asian, European, and South Asian) and UGR (i.e., Ugandan), for each population, the points represent the summed R^2^ value for each model, with the large square highlighting the model with the highest summed R^2^ value.

For the absolute medication adjustment approach, the best models for LDLc were similar amongst the East Asian, European and South Asian populations with the summed R^2^ values increasing until they peaked at either the 1.6 or 1.8 mmol/L adjustment threshold (Figure 3). In comparison, the African population showed an undulating distribution in the summed R^2^ values with the best model at the 1.2 mmol/L adjustment threshold (Figure 3). Furthermore, the Ugandan population showed a decreasing distribution in summed R^2^ values with the best model at the 0.3 mmol/L adjustment threshold (Figure 3). Taken at face value, the absolute adjustment model implies that statins have a substantially smaller effect on individuals of African population than the other major populations, whereas on the relative scale the medication effect appears to be consistent across populations. If statins do act relatively, the apparent lower absolute effect in the Ugandan population could be explained by the sample comprising younger individuals who have lower mean LDLc values, and hence for the same relative effect the absolute change from a lower mean will be smaller.

SBP findings were similar across populations with the best model at between the 5mmHg or 10mmHg absolute adjustment threshold (Figure 4). This is with the exception of the Ugandan population where the highest R^2^ value peaked at the 25mmHg medication reduction threshold model (Figure 4). Lastly, for DBP, findings were generally consistent across populations at the 5mmHg to 10mmHg adjustment threshold (Figure 4). Although the maximum R^2^ values in the relative model showed slightly stronger R^2^ values.

## Discussion

In this study, we aimed to evaluate the effects of adjusting for medication status when conducting genome-wide association analyses using the GEMRA approach. When performing simulations, our main findings suggested that the direct effects of adjusting for medication use and excluding medication use strengthened the effect estimates compared to the uncontrolled approach and were closer to the true effect estimates. We also observed greater statistical power compared to the ‘True effect’ model when applying the adjusting for medication approach followed by the uncontrolled approach which had the larger sample sizes. When sample sizes were reduced for the medication-excluded and medication-only approach, our results showed a large reduction in statistical power. Furthermore, our simulations illustrated that the indirect effects of approximately adjusting for medication status resulted in low false positive results (similar to the true effect) and results for this method were shown to, in part, resolve the collider bias issue, although some bias remains. Our simulations also showed that bias due to mismatched medication adjustment could give rise to unbiased effect estimates by applying absolute or relative medication adjustments to the unadjusted phenotype, and this bias was not largely influenced by variations in medication effect amongst individuals. Moreover, we found that overall our GEMRA approach outperformed the unadjusted model even when medication was fitted as a covariate. The GEMRA approach selected the maximum *R*^2^ value 84% of the time for a relative adjustment model compared to an absolute adjustment model when we simulated a true relative mode of medication effect.

In addition, we used real-world data to evaluate the best model (determined by the largest summed R^2^ value) when applying different thresholds to adjust observed phenotype values affected by medication use across different populations. Overall, the best model for the relative adjustment approach was consistent across all populations for LDLc (i.e. 30-40% threshold), and 10% for both SBP and DBP.

In contrast, the best model in the absolute adjustment approach was less consistent for LDLc with only the East Asian, European and South Asian populations showing similar thresholds as the best model (1.6-1.8 mmol/L threshold). For the African and Ugandan population, the best models were attenuated with an adjustment threshold of 1.2 mmol/L and 0.3 mmol/L, respectively. In contrast, the best model was similar across populations for DBP (i.e. 5-10mmHg), as well as SBP (i.e. 5-10mmHg) with the exception of the Ugandan population where the maximum *R*^2^ value peaked at 25mmHg. However, maximum *R*^2^ value for UGR (i.e. 0.0959) was very similar to the 10-15mmHg thresholds (*R*^2^=0.0936-0.0944).

Differences in medication adjustments across populations may be a result of the difference in age distribution of the individuals. This could have contributed to the heterogeneity in the best absolute model thresholds for LDLc as individuals of European descent in UKB had a median age of 58 years (Interquartile range (IQR): 45-71), whilst individuals in the UGR cohort had a median age of 30 years (IQR: 18-46). As a result, the mean observed LDLc in UGR would be relatively low due to the age distribution of the UGR cohort being younger. Therefore, a 40% reduction in medication effect of a small value is a smaller absolute change, compared to a 40% reduction of a larger value. This difference can also be seen when comparing the number of individuals taking LDLc-lowering medication (UKB EUR: 14%; UGR: <1%) (Table 1). In contrast, SBP and DBP showed more consistent absolute medication reduction thresholds across populations. This may be attribute to the greater degree of interquartile range (IQR) overlap across populations with the phenotypes ranging from 112-157mmHg for SBP and 68-96mmHg for DBP. For LDLc, there was no overlap in IQR between the Ugandan population (IQR: 1.5-2.5mmol/L) and the other populations which ranged from 2.7-4.1mmol/L.

In addition, the type of drug therapy prescribed to individuals may have contributed to the heterogeneity of best models. Randomized controlled trials (RCTs) have been conducted to evaluate the efficacy of these drug regimens with studies showing varying degrees of serum LDLc reduction(27–33) and blood pressure lowering effects(34–39). In a meta-analysis of RCTs, statin treatment was associated with a 28% reduction in LDLc levels(27) which is similar to the anticipated 30-50% reduction in LDLc levels for moderate-intensity statin therapy outlined by the 2018 US blood cholesterol treatment guidelines(40). This is also comparable to the best model thresholds identified in the relative approach across populations. Furthermore, when statin therapy is used in combination with other non-statin drugs (e.g., Ezetimibe, Evacetrapib or Evolocumab), RCTs showed a decrease in LDLc levels between 0.9-2.4 mmol/L(30–32), which is within the range of best model thresholds identified when applying the absolute approach to adjust for medication status, with the exception of the Ugandan population finding. For blood pressure, when firstly considering different antihypertensive monotherapies, an RCT meta-analysis showed SBP was reduced by 6.8-9.3 mmHg(33), which is similar to a larger meta-analysis of 208 RCTs that showed that SBP was reduced by 10-15mmHg and 8-10mmHg for DBP(33). This range is similar to our best absolute medication reduction thresholds for SBP and DBP. Interestingly, all antihypertensive drug RCTs found presented their findings using absolute medication reduction. However, our findings showed that the relative mode of medication effect was consistent across all populations.

A limitation of this study was that UKB and UGR did not report the dose, duration of treatment and level of compliance towards the medication treatments and therefore the GEMRA approach is liable to over- or under adjusting for medication effect without this additional context. As a result, some heterogeneity can be introduced into the analyses.

In addition, this study was limited by the sample size of the data sources applied in the empirical analysis. The non-European demographic groups in UKB were limited by their sample size compared to the European group (Non-European: N < 6,200; Europeans: N = 219,151). As a result, the statistical power to detect genetic variants associated with our phenotypes of interest in these groups, and subsequently to evaluate the best model using the summed R^2^ values, may have affected the precision of the findings. This can be observed when using absolute values to adjust for statin use (Figure 3B), which suggested that the average statin reduces LDLc to a greater extent in East Asians than in Europeans. However, this finding is likely a result of imprecision due to large confidence intervals surrounding the R^2^ values, as the mean population levels of LDLc tend to be notably lower in East Asian populations than in European populations. We also note that maximising R^2^ values is a convenient approach but could introduce data leakage, e.g. exacerbating winner’s curse bias in the final GWAS results. This could be mitigated by only performing GEMRA in a random subset or hold-out sample to calibrate the adjustment parameter, or by using a replication sample or winner’s curse adjustment methods post-GWAS(41).

Furthermore, although we evaluated the issue of data leakage through our simulations and real-world data examples and found no strong evidence of data leakage (Supplementary figure 6 and Supplementary figure 7) this issue could still remain a concern to researchers applying our method. Approaches similar to Sinnott-Armstrong et al. 2021(42) could be applied to mitigate this issue by estimating the adjustment factor identified used GEMRA in an independent sample, fix the selected adjustment factor, and apply it to a non-overlapping sample.

Overall, our study has illustrated the technical challenges that can arise when analysing phenotypes altered by medication status in genome-wide association analyses, and we have proposed the GEMRA approach to approximately adjust the observed values using LDLc and statin use, as well as blood pressure traits and antihypertensive use. Findings using this approach suggest that statins have a relative effect of 40% on LDLc values, and that antihypertensive drugs have a relative effect of 10% on blood pressure values which is consistent across populations. We believe that the GEMRA approach is a useful method to heuristically search for empirical adjustments to then be applied to observed phenotype values of individuals noting medication use. This method can also be broadened to allow researchers the flexibility to apply this approach to other contexts such as medications that increase the phenotype, or in contexts where a combination of the adjustment strategies is appropriate to adjust the observed phenotype value.

## Methods

### Simulation study

To assess whether medication adjustment improves the uncontrolled model to be closer to true value of the unobserved phenotype effect estimates, we conducted several simulations under different conditions. We used a basic data generating model and the observed phenotype effect estimates were estimated using a relative or absolute model as shown in the Supplementary Materials.

In the main simulation, we implemented a simulation study on 100,000 individuals with genetic effects associated with LDLc (G_direct_) and genetic effects associated with non-LDLc factors (G_indirect_) (Figure 1). We defined G_direct_ as the causal effects of genetic variants associated with LDLc on the underlying, unmedicated LDLc phenotype independent of medication status or selection factors and G_indirect_ as the apparent genetic effects arising from conditioning on medication status which induces associations between genetic variants and non-genetic factors of LDLc (i.e. collider bias) (Figure 1). The LDLc phenotype distribution was modelled on measured LDLc values in UK Biobank (UKB) with a phenotypic mean of 3.55 mmol/L and a standard deviation of 0.87 mmol/L. Furthermore, we applied a medication use prevalence of 14%, as previously reported in UKB(43), and a medication effect of 0.8 (i.e., a relative LDL-lowering medication reduction of 20%). We then repeated the simulations 1,000 times with parameters of the direct and indirect genetic effects on LDLc and the beta effect estimate for LDLc resampled in each simulation. In each simulation, we applied and compared several approaches: 1) True effect; 2) Uncontrolled approach, which made no adjustment for medication status; 3) Medication only approach, which was restricted on individuals taking statin medication; 4) Medication excluded approach, which was restricted to individuals not taking any statin medication; 5) Medication status as a covariate approach, which was restricted to unmedicated individuals or all individuals without adjustment; 6) Approximate medication adjustment approach using the GEMRA method. For the direct effect, we evaluated statistical power as the percentage difference in mean F-statistic for each method compared to the mean F-statistic for the ‘True effect’ model.

In addition, we also conducted simulations under the following conditions: 1) Incorrect adjustment for medication; 2) Variation in medication effects amongst individuals; 3) Impact of variation in medication effects on the exposure variable; 4) Robustness of the R^2^ approach to identifying the optimal medication adjustment parameter. These simulations are presented in the Supplementary Materials.

### Empirical analysis: UK Biobank and Ugandan Genome Resource

#### Cohort profiles

UKB is a population-based health research resource consisting of approximately 500,000 people, aged between 38 years and 73 years, who were recruited between the years 2006 and 2010 from across the UK(44). Particularly focused on identifying determinants of human diseases in middle aged and older individuals, participants provided a range of information (such as demographics, health status, lifestyle measures, cognitive testing, personality self-report, and physical and mental health measures) via questionnaires and interviews; anthropometric measures, blood pressure readings and samples of blood, urine and saliva were also taken (data available at www.ukbiobank.ac.uk). A full description of the study design, participants and quality control (QC) methods have been described in detail previously(45). UK Biobank received ethical approval from the Research Ethics Committee (REC reference for UK Biobank is 11/NW/0382).

The Ugandan Genome Resource (UGR) is a population-based cohort which comprises of approximately 22,000 individuals from 12 ethno-linguistic groups recruited from rural south-western Uganda(46). The initial cohort, the Ugandan General Population Cohort (GPC), was created with the purpose to study human immunodeficiency viruses in the general population. However, since 2011, this cohort has extended its scope with a subset of participants becoming the UGR cohort to also focus on understanding the epidemiology and genetics of communicable and non-communicable diseases with a wide range of phenotypic data collected (such as anthropometric measurements, demographic and lifestyle measurements through self-reported questionnaires, as well as blood specimens)(46). The Ugandan Genome Resource was approved by the Science and Ethics Committee of the Uganda Virus Research Institute (UVRI) Research and Ethics Committee (UVRI-REC #HS 1978), the Uganda National Council for Science and Technology (UNCST #SS 4283), and the East of England-Cambridge South (formerly Cambridgeshire 4) National Health Service (NHS) Research Ethics Committee UK.

#### Genotyping, imputation and quality control

A full description of the genotype generation, quality control and imputation methods have been previously reported in detail for UKB(47) and UGR(46).

In brief, for UKB, 488,377 participants were successfully genotyped with 438,427 participants genotyped using the Affymetrix UK Biobank Axiom Array and 49,950 participants genotyped using the UK BiLEVE Axiom Array. Multiallelic single-nucleotide polymorphisms (SNPs), SNPs with a minor allele frequency less than or equal to 1%, and samples that were classified as outliers for missing rate and heterozygosity were removed prior to phasing. Phasing was conducted using a modified version of the SHAPEIT2 algorithm(48), and genotype imputation to a reference set consisting of the Haplotype Reference Consortium reference panel merged with the UK10K and 1000 Genomes phase 3 reference panels(49) was performed using IMPUTE2 algorithms(50). Continental ancestry groupings were then applied using principal component and K-means clustering analyses(51).

For UGR, the 3.5M Illumina chip array was used to genotype 5,000 participants at the Wellcome Trust Sanger Institute with around 2,000 participants from the Ugandan GPC whole-genome sequenced using the Illumina HiSeq 2000(46). Pre-phasing protocols removed samples due to sex mismatch, high relatedness, call rate, and heterozygosity. Phasing was implemented using SHAPEIT2(52). Genotype imputation was conducted using IMPUTE2(53) with a reference set combining Uganda 2000 Genomes reference panel and the 1000 Genomes phase 3 reference panel(46).

#### Phenotype data

In UKB, venous blood samples were collected from participants in a non-fasting state at their baseline clinical visit between 2006 and 2010. Circulating LDLc was directly measured using an enzymatic selective protection method on a Beckman Coulter AU5800. Further details of the phenotype collection and analysis have been previously reported (https://biobank.ndph.ox.ac.uk/showcase/ukb/docs/serum_biochemistry.pdf). The blood pressure traits, SBP and DBP, were measured by automated readings using a Omron 705 IT electronic blood pressure monitor. Comprehensive details regarding the protocols followed to measure blood pressure can be found elsewhere (https://biobank.ndph.ox.ac.uk/showcase/ukb/docs/Bloodpressure.pdf). In addition, medication data (Data Field: 20003; Data coding 4) were collected through verbal interviews conducted by trained members of staff during the baseline assessment visit and subsequently during follow-up visits. Duration, dosage and compliance of medication treatments were not recorded.

In UGR, participants provided venous blood samples in a non-fasting state with circulating LDLc measured using homogenous enzymatic colorimetric assays on a Cobas Integra 400 Plus Chemistry analyser (Roche Diagnostics)(54). Protocol describing the biochemical analyses has been previous reported elsewhere(55, 56). Blood pressure measurement in UGR has been described in detail elsewhere by Gurdasani et al. 2019(54). Briefly, blood pressure was measured using the fully automated Omron M6-I.(57–59). Prior to taking measurements, participants were asked to refrain from eating and/or drinking 30 minutes before. They were also asked to rest for 15 minutes before measurements were taken. Further, the arm circumference was determined and the appropriate Omron cut-size used before measuring blood pressure. Blood pressure was measured in the sitting position three times with resting intervals of 3-5 minutes. Blood pressure for a participant was calculated as the mean of the second and third reading. Medication data for UGR was obtained from participants during enrolment through an interview. The participants would be asked if they had taken any medication for high cholesterol as prescribed by a professional health worker in the last two weeks. The responses would then be coded as per study interview codes.

#### GEMRA

We developed the genetic empirical medication reduction adjustment (GEMRA) approach to adjust the observed phenotype value in participants taking medication and restoring it to a phenotype value that is more reflective of the underlying, unmedicated value. This approach applies medication reduction adjustments (i.e., absolute and relative) to the observed phenotypic values of medicated participants. Genome-wide association analyses is conducted using unmedicated and medicated participants adjusted for medication status for each model, and R^2^ is then calculated as a measure of goodness of fit for each independent, genome-wide SNP identified in the model. R^2^ is calculated by: 1) Calculating the t-statistic using the equation, t = β/β_SE_, where β is the beta effect estimate of the SNP and β_SE_ is the standard error of the beta effect estimate of the SNP; 2) Then calculating R^2^ for each SNP using the equation, R^2^ = t^2^/(t^2^ + (n+1)), where n is the sample size. The model is determined to be the ‘best’ model for adjusting for medication effect on a phenotype of interest if the summed R^2^ values across all independent SNPs, discovered at a *p*-value < 5×10^-8^ and clumped at a r^2^ threshold of 0.1 and a distance of 250kB, is the highest.

#### Statistical analyses

GWAS were performed in UGR and all populations in UKB using a population-specific approach. GWAS was conducted using a linear mixed model association method in BOLT-LMM(v2.3)(60) for participants of European population in UKB and GENESIS(v3.17)(61) for all other populations and cohorts. All analyses were conducted using an additive genetic model and were adjusted for age (defined as the year of the survey minus participant year of birth), sex, and the first 20 principal components. Analyses were limited to genetic variants that were genotyped and imputed with a minor allele frequency >0.1% and an INFO score >0.8.

To evaluate the performance of each model, we firstly filtered the genetic variants in the GWAS output using the genome-wide threshold (*p* < 5×10^-8^) and conducted linkage disequilibrium (LD) clumping (r^2^ < 0.001) to identify independent genetic variants for each model using PLINK(v1.90) (www.cog-genomics.org/plink/1.9/)(62). We then determined the best model by calculating the R^2^ for each SNP as a metric for goodness of fit for the model. Next, we summed the R^2^ values for each model to evaluate which model was the ‘best’ by identifying the model with the highest summed R^2^ value as shown in the Supplementary Materials.

## Acknowledgements

Quality control filtering of the UK Biobank data was conducted by R.Mitchell, G.Hemani, T.Dudding, L.Corbin, S.Harrison, L.Paternoster as described in the published protocol (doi: 10.5523/bris.1ovaau5sxunp2cv8rcy88688v).

## Funding

This research has been conducted using the UK Biobank Resource under Application No 15825. This project was supported by the UK Medical Research Council (MRC) as part of a network grant awarded to the MRC Integrative Epidemiology Unit at the University of Bristol (MC_UU_00011/4, MC_UU_00011/1, MC_UU_00032/1, MC_UU_00032/3), core funding to the MRC/Uganda Virus Research Institute and London School of Hygiene & Tropical Medicine (LSHTM) Uganda Research Unit under the MRC/UK Department of International Development (DFID) Concordat agreement with Segun Fatumo supported by the Wellcome Trust (220740/Z/20/Z), and the MRC Population Health Research Unit at the University of Oxford (MC_UU_00017/2).

This study was also supported by the NIHR Biomedical Research Centre at the University Hospitals Bristol and Weston NHS Foundation Trust and the University of Bristol. This publication is the work of the authors who will serve as guarantors for the contents of this paper. The views expressed in this publication are those of the author(s) and not necessarily those of the NHS, the National Institute for Health Research.

## Data availability

UK Biobank data used in this study was obtained upon approval of the project application (Research application number: 15825). UK Biobank data is available to approved researchers upon application through the UK Biobank Access Management System (https://www.ukbiobank.ac.uk/use-our-data/apply-for-access/). For the Ugandan Genome Resource, data used in this study is available upon request following the MRC/UVRI and LSHTM Uganda Research Unit’s data sharing policy which can be found at: https://apps.mrcuganda.org/mrcdatavisibility/Home/. Requests for access to the genetic data can be directed to.

## Code availability

All scripts used to perform these simulations and analyses are available at https://github.com/MRCIEU/medication-adjustment.

## Supporting information

**Supplementary figure 1. Evaluation of bias in genetic effect estimates under unadjusted, absolute adjustment, and relative adjustment approaches.** The use of unadjusted observed values of *y_obs_* as the outcome in the model lead to bias in both *β̂_y_* and *b̂_y_* Over or under adjustment using absolute adjustment (A) or relative adjustment (B) can lead to worse bias in these estimates than the unadjusted model, though absolute and relative adjustments can both give rise to unbiased effect estimates.

**Supplementary figure 2. Impact of heterogeneity in medication effects on genetic effect estimates under relative adjustment.** We observed that this pattern of bias was not influenced by introducing heterogeneity into medication effects, where *σ*^2^ = (0, 0.1, 0.2), though standard errors increased as heterogeneity increased.

**Supplementary figure 3. Comparison of absolute and relative adjustment approaches for identifying optimal medication adjustment parameters.** A) The true medication effect operates on the relative scale in our simulations. Over the range of adjustment parameters both absolute and relative adjustment show improvements in *R*^2^(*g*, *y*) over unadjusted values of *y_obs_*, however the relative model outperforms the absolute model. Note that the relative adjustment performs best when *γ*^∗^ (x-axis) is close to the true value of *γ* = 0.8. B) We show also that selecting the best model (e.g. maximising for *R*^2^(*g*, *y*)) selects a value of *γ^∗^* for the relative adjustment that gives an estimate of *β̂_y_* that is close to the true estimate of *β_y_* = 1.

**Supplementary figure 4. Evaluation of the performance of the maximum R^2^ approach to identify the optimal adjustment model, where model operates on a relative scale in these simulations.**

Supplementary figure 5. Comparison of the mean squared error (MSE) of the effect estimate using the unadjusted phenotype (y-axis) against the adjusted phenotype (x-axis), where the adjusted phenotype is based on the adjustment factor that maximises *R*^2^(*g*, *y*).

**Supplementary figure 6. Comparison of simulated beta estimates comparing the ‘True effect’ model and the approximate adjustment for medication model.**

**Supplementary figure 7. Comparison of European LDLc GWAS beta effect estimates using population-specific absolute adjustment thresholds.** Absolute adjustment factors (*δ*\*) were estimated using the optimal absolute adjustment threshold (i.e. threshold yielding the largest maximum summed R^2^) identified in each independent population: 0.3 mmol/L in Ugandan, 1.6 mmol/L in African, and 1.8 mmol/L in East Asian populations. These adjustment factors were then applied to a non-overlapping European sample and compared with beta effect estimates obtained using the optimal 1.6 mmol/L threshold identified in the European overlapping sample. Each point represents a beta effect estimate under the corresponding population-specific threshold.

## Notes

### Competing Interest Statement

The authors have declared no competing interest.

### Author Declarations

This project used cohort data that received ethical approval. UK Biobank received ethical approval from the Research Ethics Committee (REC reference for UK Biobank is 11/NW/0382). The Ugandan Genome Resource was approved by the Science and Ethics Committee of the UVRI Research and Ethics Committee (UVRI-REC #HS 1978), the Uganda National Council for Science and Technology (UNCST #SS 4283), and the East of England-Cambridge South (formerly Cambridgeshire 4) NHS Research Ethics Committee UK.

### Summary of Updates

Main manuscript revised; Supplementary material file updated.

