## Supplementary Information for "Adjusting for medication status in genome-wide association studies"

### Supporting information

**Data generating model**

The basic data generating model is as follows. The true biological phenotype (i.e. that has not been adjusted by medication) is simply

$$y_i = \alpha + \beta_y g_{y,i} + \epsilon_i$$

Where $i$ represents an individual, $g_{y,i}$ is that individual's genetic value for the phenotype which could be a single SNP or a polygenic score, $\beta_y$ is the direct effect of that genetic value and $e_i \sim N(0, \sigma^2)$ is the residual variance. We fix $\alpha = 130$ and $\sigma = 20$ to reflect the distribution of LDL cholesterol in UK Biobank. Individuals have a liability for taking medication

$$l_i = (y_i - \alpha)/\sigma + \beta_m g_{m,i}$$

where $g_{m,i}$ is the genetic value for individual $i$ for taking medication that is independent of $g_{y,i}$, and $\beta_m$ is the effect of the medication genetic liability. Therefore individuals have probability of medication based on

$$\rho_i = 1/\left(1 + e^{l_i}\right)$$

and medication status is sampled based on

$$m_i \sim Binom(1, p_i)$$

The effect of medication on individuals for whom $m = 1$ depends on the true mode of effect of the medication. For a relative effect the observed phenotypic value after medication is

$$y_{obs,(m=1)} = y_{(m=1)}(\gamma)$$

where $\gamma$ is the relative effect of medication. Alternatively, an absolute effect on medication would be

$$yobs,(m{=}1) = y(m{=}1) + \delta$$

Where $\delta$ is the absolute effect of medication. Note that the medication effect can be heterogeneous across individuals such that $\gamma_i \sim N(\gamma, \sigma_\gamma^2)$.

Because the true value of $y_i$ is unobserved, estimation of the direct effect $\beta_y$ is obtained from

$$yobs*,i = \hat{\alpha} + \beta ygy,i + \epsilon i$$

where $y_{obs^*,i}$ is the observed phenotype that has potentially been adjusted.

An indirect effect ($b_y$) can be induced when sample selection is based on medication status (i.e., conditioning on medication status through the inclusion or exclusion of participants, or stratifying by medication status) (Figure 1). This opens up a causal pathway through medication status to the observed phenotype value as

$$y_{obs^*,i} = \hat{\alpha} + \beta ygy,i + b_y g_{y,i} + \epsilon i$$

Giving rise to collider bias, and biasing the causal effect estimate ($y_{obs^*,i}$).

Adjustment is performed using either a relative model whereby an adjustment factor $\gamma^*$ is selected such that

$$yobs^*,(m{=}1) = yobs,(m{=}1)\gamma^*$$

or an absolute model whereby an adjustment factor $\delta^*$ is selected such that

$$y_{obs^*},(m{=}1) = yobs,(m{=}1) + \delta_*$$

We also estimate the effect of $g_m$ on $y_{obs}$, which should be null but an effect can be induced due to collider bias arising from medication:

$$yobs*,i = \hat{a} + \hat{b}ygm,i + ei$$

**Model adjustment parameter selection**

To select the 'best' model, we first calculated the *t*-value for each independent SNP in each model by

$$t = \frac{\beta}{SE}$$

We then calculate the $R^2$ value for each independent SNP, where *M* represents the number of independent significant SNPs in each analysis and *j* represents the t-statistic for SNP $j$, and sum the $R^2$ values such that

$$R^2_{model} = \sum_{j=1}^{M} \frac{t_j^2}{t_j^2 + (n-1)}$$

**Evaluation of statistical power**

Statistical power was evaluated as the percentage difference in the mean F-statistic for each method compared to the mean F-statistic for the 'True effect' model. This method was applied to evaluate the loss in power of the methods (i.e. Uncontrolled, medication only, medication excluded, medication status as a covariate and approximate adjustment for medication) compared to the 'True effect' model. For each independent SNP used in the models the *F* statistic was calculated by

$$F = \frac{\beta^2}{SE^2}$$

Difference in statistical power was then calculated as the percentage difference compared to the 'True model' was then obtained by

$$Difference\ in\ power\ =\ \frac{\bar{F}_{method}}{\bar{F}_{true}} \times\ 100$$

We used this approach to evaluate power rather than converting F to a probability of discovering a true positive rate to avoid the need to choose simulation parameters that were liable to flattening out (e.g. at high or low power settings).

**Simulation parameters**

To evaluate the impact of incorrectly adjusting for medication we performed a range of simulations whereby the sample size used in the data generation model was 100,000, $\beta_y = 1$, $\beta_m = 1$, $\sigma_y{}^2 = 0$ and for simplicity we simulated $g_m$ and $g_y \sim N(0,1)$. We varied the values of $\gamma^* = (0.1,0.2, \ldots ,1.4)$ and $\delta^* = (0,10,20, \ldots ,80)$. The mode of medication effect on $y$ is always relative in these simulations such that $\gamma = 0.8$ and $\delta = 0$. Each simulation was performed 100 times and the effect estimate of $\hat{\beta}_y$ and $\hat{\beta}_m$ was obtained on the true value $y$, unadjusted $y_{obs}$, and relative and absolute adjusted values of $y_{obs}{}^*$. Findings from this simulation are shown in Supplementary figure 1 and 2.

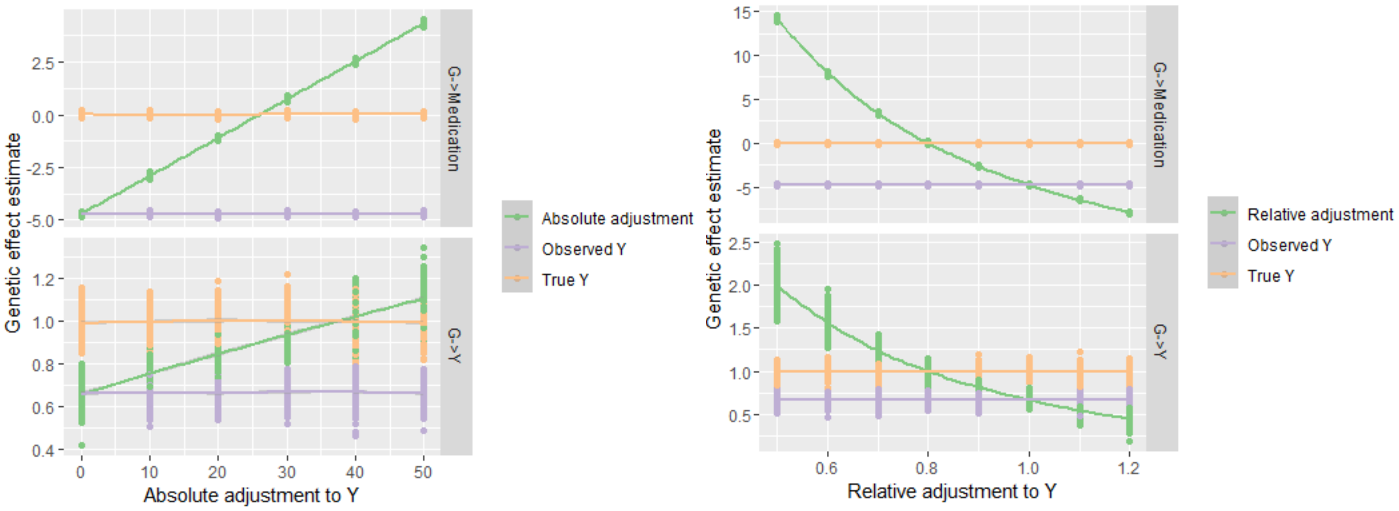


Supplementary figure 1. The use of unadjusted observed values of $y_{obs}$ as the outcome in the model lead to bias in both $\hat{\beta}_y$ and $\hat{b}_y$. Over or under adjustment using absolute adjustment (A) or relative adjustment (B) can lead to worse bias in these estimates than the unadjusted model, though absolute and relative adjustments can both give rise to unbiased effect estimates.

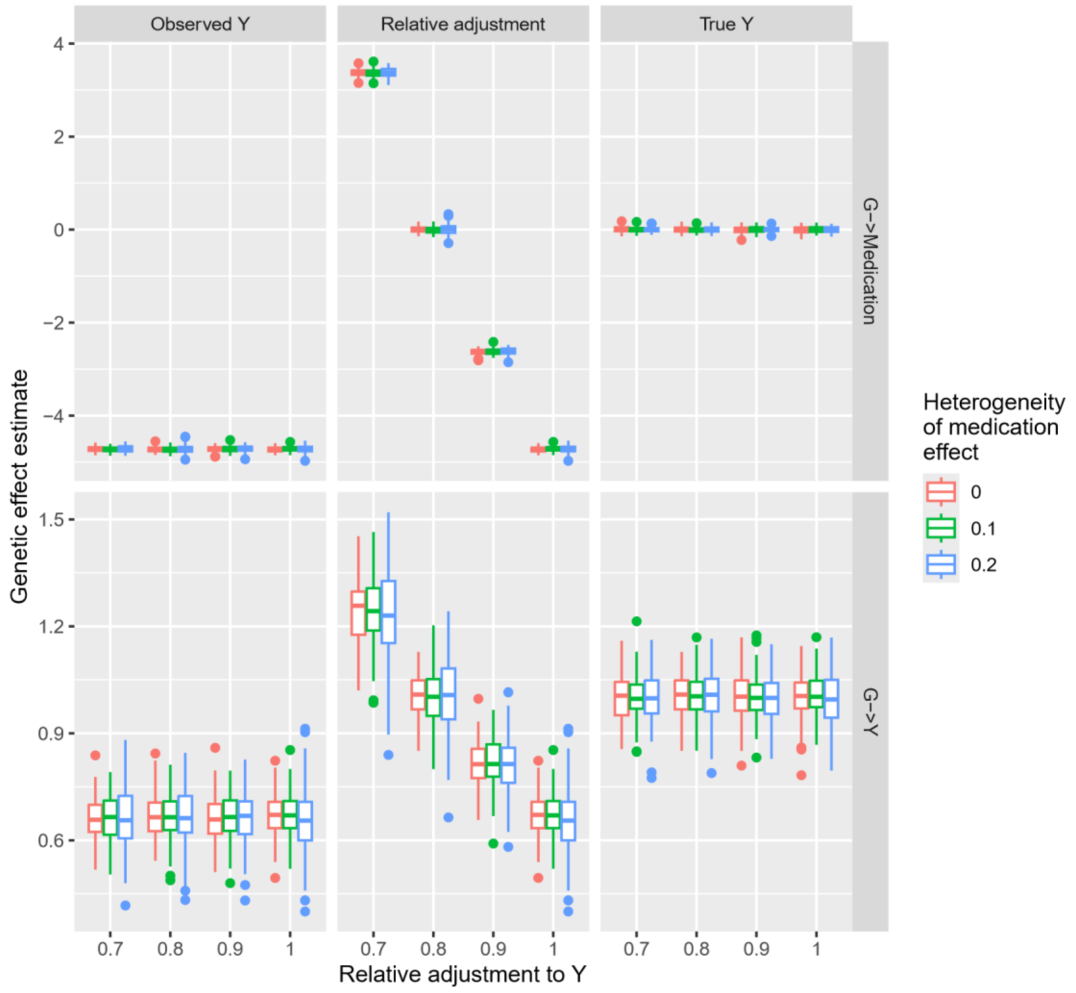


Supplementary figure 2. We observed that this pattern of bias was not influenced by introducing heterogeneity into medication effects, where $\sigma_\gamma^2 = (0, 0.1, 0.2)$, though standard errors increased as heterogeneity increased.

**Detection of adjustment parameters**

In order to identify the adjustment parameter that minimises bias in GWAS, we searched over a range of adjustment parameters using known genetic variants, seeking the adjustment parameter that maximises $R^2(g, y)$. An example output from a single simulation is shown in Supplementary figure 3.

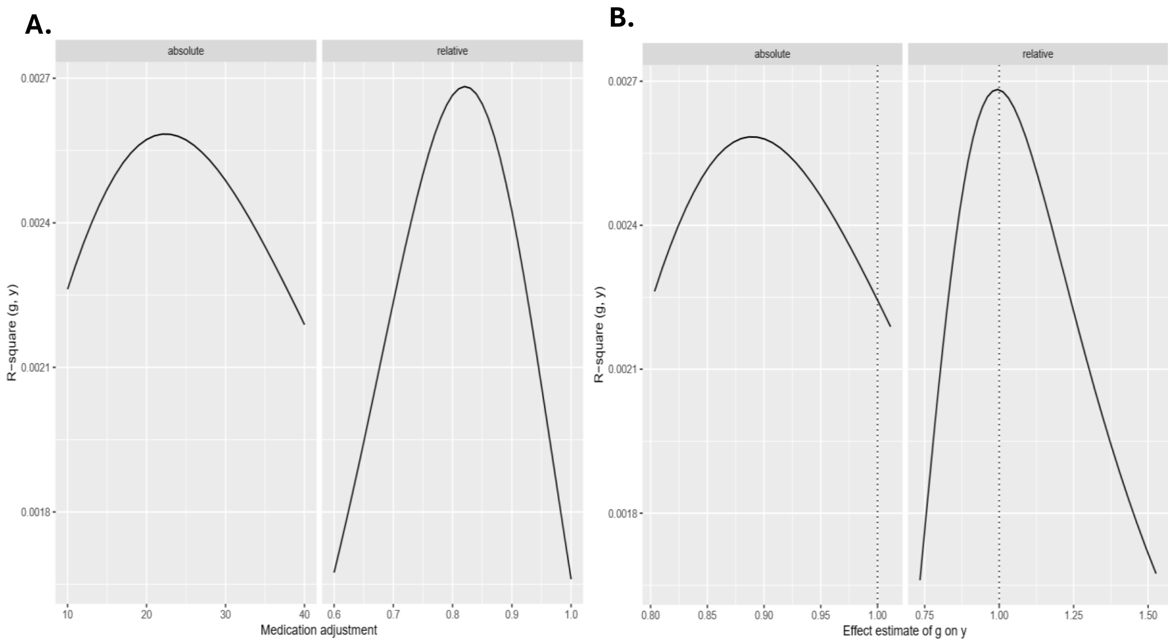


Supplementary figure 3. A) The true medication effect operates on the relative scale in our simulations. Over the range of adjustment parameters both absolute and relative adjustment show improvements in $R^2(g, y)$ over unadjusted values of $y_{obs}$, however the relative model outperforms the absolute model. Note that the relative adjustment performs best when $\gamma^*$ (x-axis) is close to the true value of $\gamma = 0.8$. B) We show also that selecting the best model (e.g. maximising for $R^2(g, y)$) selects a value of $\gamma^*$ for the relative adjustment that gives an estimate of $\beta_y$ that is close to the true estimate of $\beta_y = 1$.

We performed 1,000 repeats of these simulations to evaluate the performance of this method and show that 84% of the time the value of $R^2(g, y)$ is maximised for a relative adjustment model vs an absolute adjustment model (Supplementary figure 4).

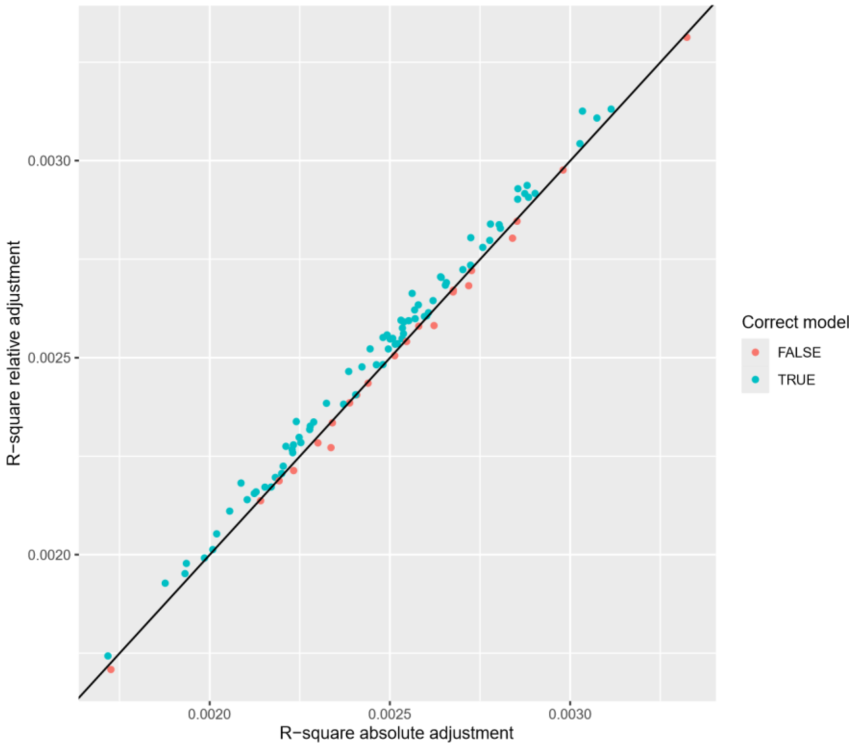


Supplementary figure 4. Evaluation of the performance of the maximum $R^2$ approach to identify the optimal adjustment model, where model operates on a relative scale in these simulations. Finally, in Supplementary figure 5, though in 16% of cases the adjustment factor uses an absolute rather than a relative model, adjustment is always outperforming unadjusted analysis (A). The adjustment strategy also outperforms using the unadjusted phenotype with medication being fitted as a covariate in almost all simulations also (B).

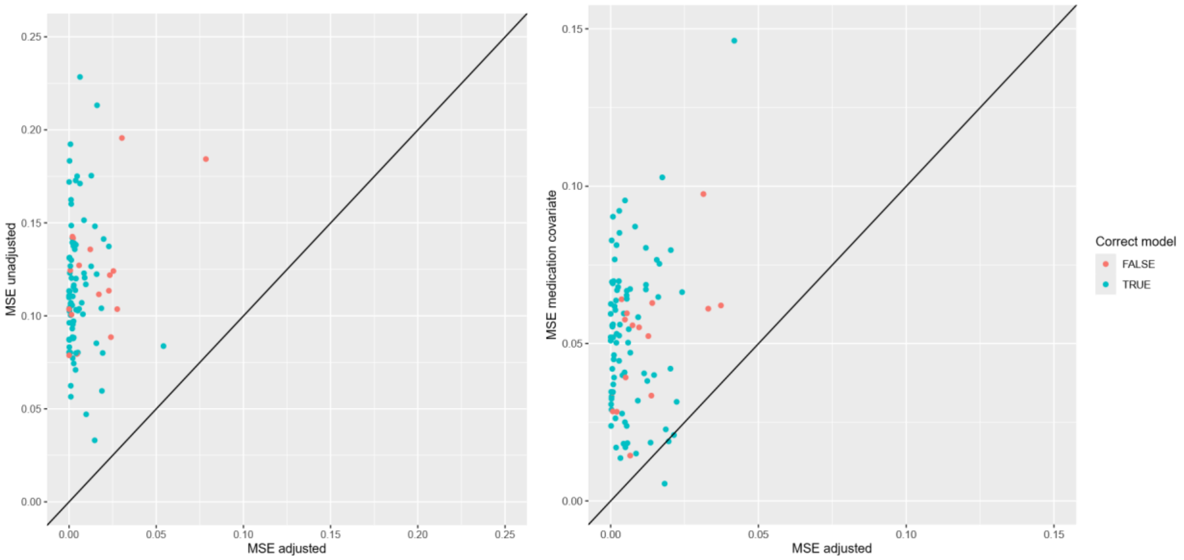

Supplementary figure 5. Comparison of the mean squared error (MSE) of the effect estimate using the unadjusted phenotype (y-axis) against the adjusted phenotype (x-axis), where the adjusted phenotype is based on the adjustment factor that maximises $R^2(g, y)$.

**Evaluation of data leakage**

We evaluated the issue of data leakage by comparing the direct genetic effect estimates using the GEMRA approach (i.e. Approximate adjustment for medication) and the 'True effect' model, and we found no evidence of bias due to data leakage with findings showing a strong, positive linear correlation (Supplementary figure 6). We then applied a similar approach used by Sinnott-Armstrong et al. using the absolute adjustment thresholds identified as the best model (i.e. threshold with the largest maximum summed $R^2$ value) for each population in our LDLc real-world data GWAS findings and applied them to our European dataset. This approach would estimate the absolute adjustment factor ($\delta^*$) in an independent sample (i.e. African, East Asian and Ugandan population) and apply it to a non-overlapping sample (European population). In Supplementary Figure 7, we compared the European beta effect estimates for the 0.3 mmol/L, 1.6 mmol/L and 1.8 mmol/L identified as the best model from the African, East Asian and Ugandan populations, respectively, compared to the 1.6 mmol/L model in Europeans as the overlapping sample. Our findings showed no strong evidence of data leakage with similar beta effect estimates in the non-overlapping samples compared to the overlapping European sample.

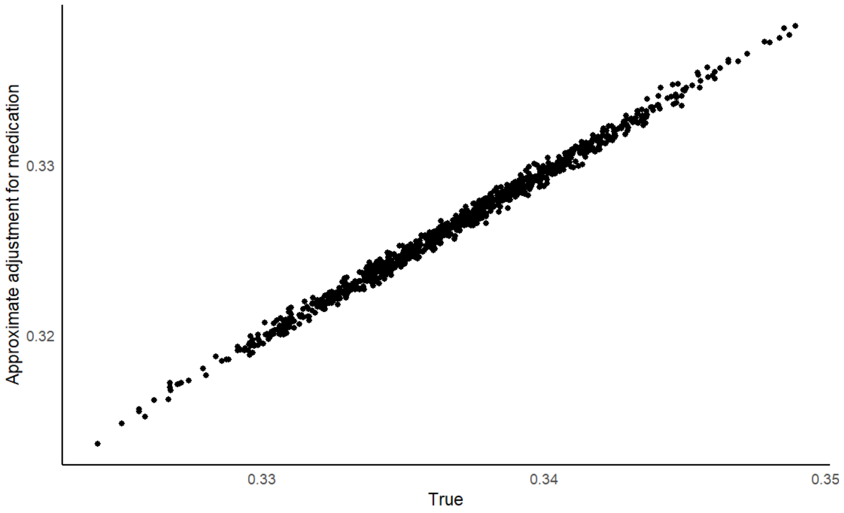

Supplementary figure 6. Comparison of simulated beta estimates comparing the ‘True effect’ model and the approximate adjustment for medication model.

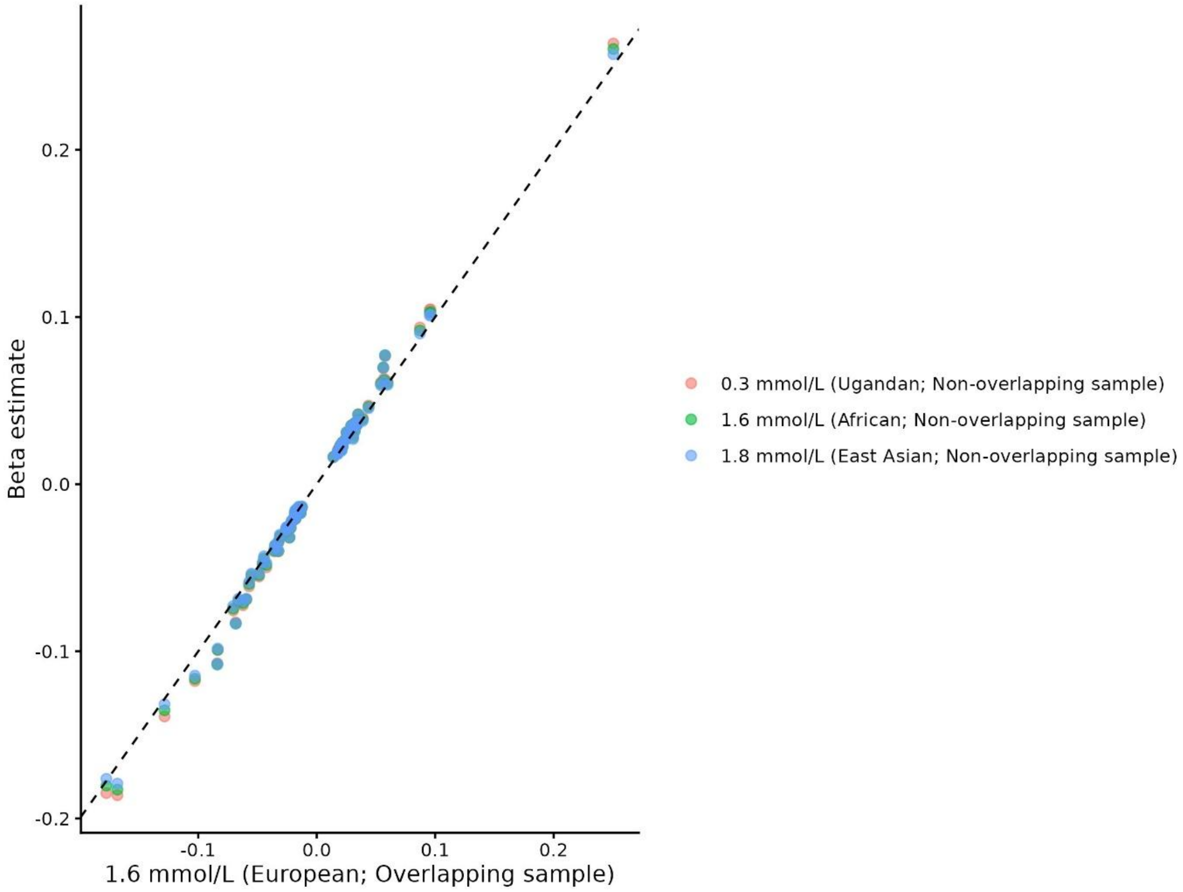


Supplementary figure 7. Comparison of European LDLc GWAS beta effect estimates using population-specific absolute adjustment thresholds. Absolute adjustment factors ($\delta^*$) were estimated using the optimal absolute adjustment threshold (i.e. threshold yielding the largest maximum summed $R^2$) identified in each independent population: 0.3 mmol/L in Ugandan, 1.6 mmol/L in African, and 1.8 mmol/L in East Asian populations. These adjustment factors were then applied to a non-overlapping European sample and compared with beta effect estimates obtained using the optimal 1.6 mmol/L threshold identified in the European overlapping sample. Each point represents a beta effect estimate under the corresponding population-specific threshold.
